# Patients Recovering from COVID-19 who Presented Anosmia During their Acute Episode have Behavioral, Functional, and Structural Brain Alterations

**DOI:** 10.1101/2024.05.01.24306655

**Authors:** Leonie Kausel, Alejandra Figueroa-Vargas, Francisco Zamorano, Ximena Stecher, Mauricio Aspé-Sánchez, Patricio Carvajal-Paredes, Victor Márquez-Rodríguez, Maria Paz Martinez-Molina, Claudio Román, Patricio Soto-Fernández, Gabriela Valdebenito-Oyarzo, Carla Manterola, Reinaldo Uribe, Claudio Silva, Rodrigo Henríquez-Ch, Francisco Aboitiz, Rafael Polania, Pamela Guevara, Paula Muñoz-Venturelli, Patricia Soto-Icaza, Pablo Billeke

## Abstract

Patients recovering from COVID-19 commonly exhibit cognitive and brain alterations, yet the specific neuropathological mechanisms and risk factors that underlie these alterations remain elusive. Given the significant global incidence of COVID-19, identifying factors that can distinguish individuals at risk of developing medium or long-term brain alterations is crucial for prioritizing follow-up care. Here, we report findings from a sample of 100 patients who were affected by a respiratory infection during the COVID-19 pandemic. This sample comprised 73 adults with a mild to moderate SARS-CoV-2 infection (who did not require invasive ventilatory assistance) and 27 with infections attributed to other agents and no history of COVID-19. The participants underwent cognitive screening, a decision-making task to measure cognitive flexibility, and magnetic resonance imaging evaluations. We assessed two clinical factors during infection: the presence of anosmia and the requirement for hospitalization due to respiratory symptoms. Groups did not differ in age or cognitive performance, but clinical factors differentially affected task performance. Patients who presented anosmia during the acute episode exhibited more impulsive changes in alternatives after a shift in probabilities in the decision-making task, while patients who required hospitalization showed more perseverative choices. Interestingly, the presence of anosmia during SARS-CoV-2 infection correlated with several brain measures, including decreases in functional activity during the decision-making task, thinning of cortical thickness in parietal regions, and loss in white matter integrity in corticospinal tracts and parietal-thalamic fasciculi, among others. These results suggest that anosmia could be a risk factor for developing brain alterations after SARS-CoV-2 infection and may serve to identify at-risk populations for follow-up.

## Introduction

Since the onset of the pandemic caused by the SARS-CoV-2 virus, evidence has accumulated that some recovered patients from the acute infection episode exhibit cognitive impairment and brain alterations.^1–3^ Studies have found that even months after recovery from COVID-19 infection, some individuals continue to experience neurological, psychiatric, and cognitive effects.^4,5^ Persistent cognitive symptoms have been linked to brain alterations, including cerebral hypoperfusion^6^ and hypometabolism.^7^ Indeed, patients recovering from non-severe COVID-19 without cognitive symptoms present cortical thickness alterations and changes in white matter integrity.^1^ Although specific brain alterations may not always be directly related to detectable cognitive changes in COVID-19 patients,^2^ a specific visuoconstructive deficit was detected in approximately one-quarter of mild COVID-19 individuals. This deficit was associated with molecular and structural brain imaging changes and correlated with the upregulation of peripheral immune markers.^3^

Despite the increasing body of evidence, the specific clinical factors associated with brain alterations remain elusive, presenting challenges in identifying populations at risk of developing long-term brain and cognitive impairments following SARS-CoV-2 infection. The severity of acute infection is the most studied clinical factor that leads to brain and cognitive alteration. A longitudinal cohort study has revealed that the severity of the acute episode correlates with cognitive impairments in long-term follow-up.^8^ Furthermore, patients who experienced a severe episode and exhibited neurological symptoms during the infection displayed cognitive impairment and brain alterations one-year post-infection.^9^ Moreover, even patients with mild COVID-19 also present subtle brain alterations.^1–3^ It remains unknown which risk factors or clinical profiles distinguish mild COVID-19 patients who may present this alteration from those who do not.

Anosmia is a commonly reported symptom in COVID-19 patients that frequently occurs early in the course of the disease and may persist as a long-term symptom.^10,11^ It has been suggested that SARS-CoV-2 might enter the central nervous system via the olfactory epithelium, causing damage to the olfactory receptor nerves and resulting in anosmia.^12^ Although the virus primarily affects non-neuronal cells in the olfactory epithelium,^13,14^ the presence of SARS-CoV-2 in nasal brushings and olfactory mucosa biopsies of COVID-19 patients further supports the notion that the virus can invade olfactory neurons following the primary involvement of non-neuronal cells.^15^ These potential mechanisms are further bolstered by evidence from animal models.^16^ However, exhaustive research has not provided categorical evidence demonstrating the virus’s direct infection of the central nervous system in humans.^17^ Additionally, the presence of anosmia during the acute episode may indicate susceptibility to brain damage through other mechanisms, as this symptom is a known risk factor for dementia and cognitive decline.^18,19^ In this line, the presence of anosmia is associated to the UGT2A1/UGT2A2 locus that encodes for an enzyme expressed in the olfactory epithelium.^20^ Interestingly, this enzyme is also expressed in the brain and other tissues, serving as a chemical barrier to protect against xenobiotics.^21^ To date, no reports have evaluated anosmia and the severity of respiratory symptoms as independent risk factors for brain alterations caused by SARS-CoV-2 infection.

Here, we assessed whether hospitalization requirement, as a proxy for respiratory symptom severity, or anosmia, is associated with behavioral and brain alterations in adults recovering from mild to moderate COVID-19. Our findings indicate that both hospitalization and anosmia have differential impacts on behavior in a cognitive flexibility task. However, only anosmia is consistently correlated with alterations in brain function across several parameters including functional activity, cortical thickness, and white matter integrity.

## Materials and methods

### Sample

We investigated a sample of 100 patients aged between 18 and 65 years at the time of participation in the study, of whom 73 were confirmed to have a respiratory infection caused by SARS-CoV-2 through PCR diagnosis. The remaining 27 patients presented with a respiratory infection attributable to an infectious agent other than SARS-CoV-2, as evidenced by at least two negative PCR tests for SARS-CoV-2 during the acute episode or subsequent periods, and no history of COVID-19 symptoms or positive PCR for SARS-CoV-2 up to the time of evaluation. All participants were asymptomatic for respiratory issues for at least four weeks before the assessments. Exclusion criteria included the need for invasive ventilatory support or admission to intensive care units, pre-existing brain lesions, neuropsychiatric disorders, or vascular injuries either before or during the COVID-19 episode, and neurological symptoms other than anosmia during acute episodes, including seizures. All participants gave their written informed consent. Experiments were conducted at the Social Neuroscience and Neuromodulation Laboratory at the Centro de Investigación en Complejidad Social (neuroCICS) at Universidad del Desarrollo and the Unidad de Imágenes Cuantitativas Avanzadas (UNICA) at Clínica Alemana de Santiago. All participants gave their informed consent, and all experimental procedures were approved by the Ethics Committee of Clínica Alemana – Universidad del Desarrollo, Chile (Folio 2020-102). The participants’ ages ranged from 19 to 65 years (median = 39, mean = 40.1), with no significant differences observed between COVID-19 and non-COVID-19 patients (t-test, t = –1.2, df = 45.6, p-value = 0.2, r = 0.12). The time elapsed between diagnosis and the initial evaluation ranged from 2 to 11 months (median = 5, mean = 4.9), with no significant differences observed between COVID-19 and non-COVID-19 patients (t-test, t = –1.4, df = 42.7, p = 0.15, r = 0.2).

### Assessment

All participants underwent assessment using the following screening tests to estimate their overall functioning: (i) Executive functioning was evaluated using the Chilean version of the Ineco Frontal Screening (IFS-Ch)^22^. This test assesses performance in response inhibition, set shifting, working memory, and abstraction capacity. (ii) The assessment of cognitive impairment was conducted using the Chilean version of the Addenbrooke’s Cognitive Examination – ACE-III^23^. This cognitive screening test consists of 5 subscales representing specific cognitive domains (orientation and attention, memory, verbal fluency, language, and visuospatial skills). (iii) The presence of anxiety symptoms was assessed using the Spanish version of the Generalized Anxiety Disorder 7-item (GAD-7).^24^ The GAD-7 is a brief self-report scale comprising 7 items designed to identify probable cases of Generalized Anxiety Disorder (GAD) based on the diagnostic criteria of the Diagnostic and Statistical Manual of Mental Disorders – fourth edition (DSM-IV). (iv) The assessment of depressive symptoms was conducted using the Spanish version of the Patient Health Questionnaire-9 (PHQ-9)^25^. The PHQ-9 consists of 9 items that assess the presence of depressive symptoms (corresponding to DSM-IV criteria) experienced in the past 2 weeks. Additionally, the olfactory function was evaluated using the rapid psychophysical olfactory (KOR) test,^26^ and the functional capacity was evaluated using the Six Minute Walk Test (6MWT).^27^ The KOR test is a screening validated for use in detecting olfactory deficits in patients with COVID-19.^26^

### Behavioral Task

The participants completed the Reversal Learning Task (RLT, Figure 1A) in two rounds during the same session: one behavioral session outside the scanner (without imaging) and one concurrent with functional magnetic resonance imaging (fMRI) acquisition. The rounds were jointly analyzed to enhance the robustness of the behavioral analysis.

**Figure 1.**
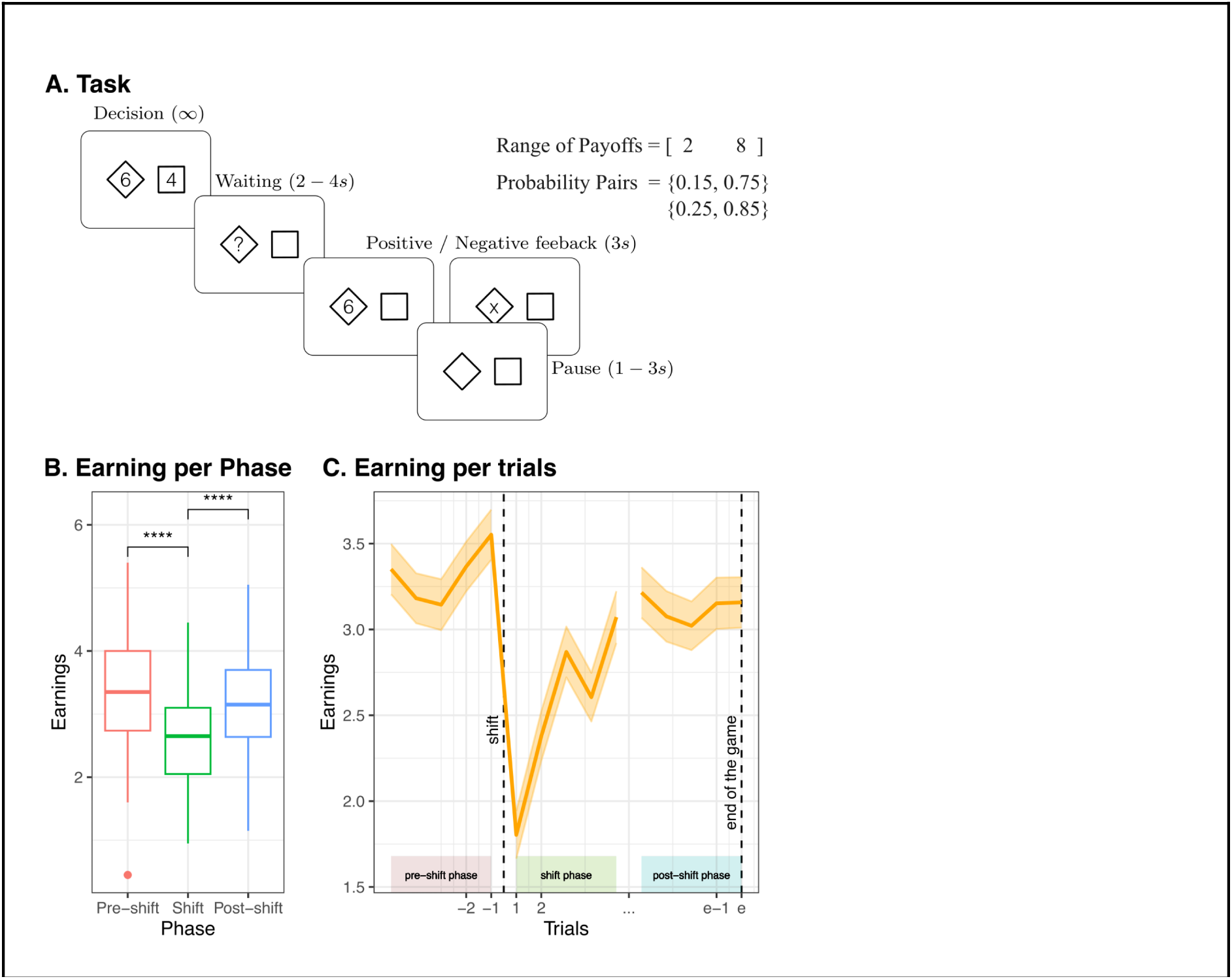
Reversal learning task. **A.** Timeline of a trial. **B.** Earnings for the three phases of the task for the complete sample. **C.** Trial means and standard errors of the evaluation of earnings during the three phases of the task for the complete sample.

In this task, participants had to choose between two decks of cards presented to them on the computer screen. In each trial, each deck was associated with a score indicating the potential winnings if that chosen deck was the paying deck for that trial. Once participants selected one of the two decks, it was revealed whether the chosen deck paid the indicated score or not. The payout probabilities for each deck were unknown to the participants, and they had to approximate them through experience. The programmed probabilities for the decks were 0.85/0.15 or 0.75/0.25. After 10 to 15 trials, the probabilities of the decks were inadvertently switched for the participant, and this arrangement remained stable for another 10 to 15 rounds until the end of the game. Each participant played four games outside the scanner and four games inside the scanner.

Participants were instructed that the goal was to accumulate the highest possible number of points and that the probability of the decks in some games might change inadvertently. The underlying logic of this task is that subjects must optimize their responses based on the scores offered and what they have learned about the payout probabilities of the decks. Additionally, during the changes, subjects must adapt and alter the learned probability for each deck to optimize their responses in these new circumstances. Therefore, the task represents an environment with high volatility that demands continuous adaptation and a balance between maintaining a stable representation of the value of the options and detecting environmental changes to adjust the values.^28^

### Cognitive modeling

The participants’ responses were analyzed using a computational cognitive methodology (see, for example,^29^). All computational cognitive models were calculated utilizing Prospect Theory, which postulates that an option’s expected subjective value or utility (U) is determined by the individuals following Equation 1.

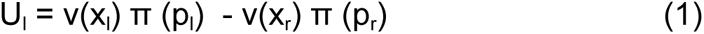

In Equation 1, subindices ‘l’ and ‘r’ represent the left and right options. v(.) represents the value function, x_l_ and x_r_ denote the potential outcome of each option associated with the left or right option, respectively. The value function was calculated using Equation 2.

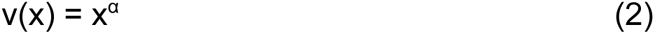

Here, α determines the concavity of the value function. ‘p’ is the probability of a gain, whereas π(.) is the subjective decision weights assigned to these probabilities. To accommodate for the learning of the unknown probabilities, the subject estimated probability by which the outcome x occurs is defined by equations 3, 4, and 5.

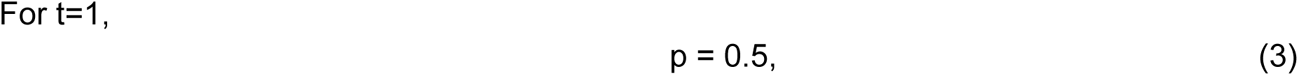

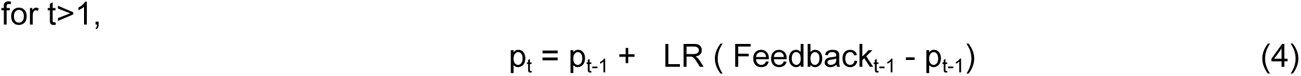

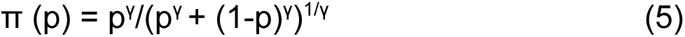

In the preceding equations, ‘γ’ is the subjective probability distortion given by Prospect-Theory, ‘t’ represents the trials during a game, and ‘LR’ represents the learning rate parameter of the Rescorla-Wagner algorithm. ‘Feedback_t-1_’ is a dummy parameter that takes the value of one if the preceding subject’s choice was rewarded and zero otherwise. We explored two variations in this algorithm: one involved fitting different learning rates (LR) following negative or positive feedback, and the other involved updating the value for only the chosen option or updating both options, assuming the opposite outcome for the other option (counterfactual updating). Since the model with different LR for positive and negative feedback and counterfactual updating provided the best data fitting, we utilized this variation for all subsequent analyses, as shown below.

The probability of choosing the left option for a given subjective value is computed using a logistic choice rule wherein β_1_ is the inverse temperature parameter representing the degree of stochasticity in the choice process, and β_0_ is a bias parameter.

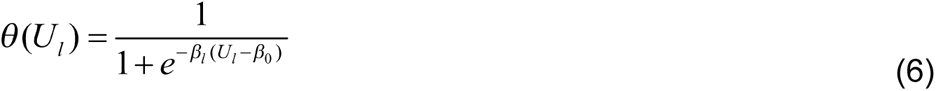

All parameters were estimated using a Bayesian approach for each individual rather than a hierarchical model due to computational limitations in using this approach in a large sample with a complex model. At the trial level, choices were modeled following a Bernoulli process:

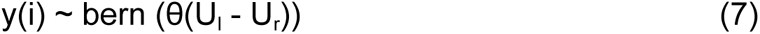

The posterior inference of parameters within the Bayesian models was executed through the Gibbs Sampler utilizing the Markov Chain Monte Carlo (MCMC) technique, implemented in JAGS via R software. An initial burn-in sequence yielded a minimum of 10,000 samples. Then, 10,000 new samples were drawn across three chains, each generated by a random number generator engine employing different seeds. In cases where convergence criteria were not met, the length of the burn-in sequence was extended. Thinning by a factor of 10 was applied to this sample set, resulting in a final collection of 3,000 samples for each parameter. Thinning was employed to mitigate autocorrelation among the ultimate parameter samples of interest. Gelman-Rubin Tests were conducted for each parameter to affirm chain convergence. The Gelman-Rubin statistic for all latent variables in our models approached 1, indicating convergence of all three chains toward the target posterior distribution.

### Imaging acquisition

All participants underwent (i) a sagittal 3D anatomical MPRAGE T1-weighted imaging (repetition time [TR]/ echo time [TE]=2530/2.19 ms, inversion time [TI]=1100 ms, flip angle=7°; 1×1×1 mm3 voxels), (ii) a sagittal 3D anatomical SPC T2-weighted (TR/TE=3200/412 ms, flip angle=120°; echo train length [ETL]=258; 1×1×1 mm3 voxels), (iii) a sagittal 3D fluid attenuation inversion recovery (FLAIR) imaging (TR/TE=5000/388 ms, TI=1800 ms, ETL=251; flip angle=120°, 1-mm slice thickness, 1×1×1 mm3 voxels), (iv) an axial 3D echo-planar imaging (EPI) (TR/TE=8600/95 ms, 2×2×2 mm3 voxels, flip angle = 90°) with diffusion gradients applied in 30 non-collinear directions and two optimized b factors (b1 = 0 and b2 = 1000 s/mm2) with three repetitions, and (v) a functional image (fMRI) weighted echo-planner T2* (TR/TE=2390/35 ms, flip angle=90°, 3 × 3 × 3 mm voxels). All imaging was acquired on a 3T Siemens Skyra (Siemens AG, Erlangen, Germany) MR scanner with a gradient of 45 mT/m and a maximum slew rate of 200 mT/m/s.

### Imaging analyses

The whole brain was acquired for fMRI imaging while the experimental task was executed. Participant volumes were coregistered to 2-mm standard imaging using the nonlinear algorithm, FNIRT, implemented in FSL.^30^ The BOLD signal was analyzed using different models, including motion correction parameters (MC). During decision-making periods, we fitted a model as follows.

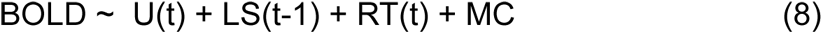

In Equation 8, U(t) represents the utility of the choice option for the current trial (t), estimated through the mean of prospect theory and adjusted using subject responses (refer to Equation 1). LS represents the learning signal from the preceding trial (t-1), derived from the unsigned prediction error of the feedback from the prior trial. By employing this regressor, we can control for signals derived from previous trials rather than the actual evaluation of the option, and it also functions as a proxy to assess the perceived uncertainty of the environment. RT represents the reaction time used as a proxy to control for the choice difficulty. During the feedback period, we fitted a model as follows.

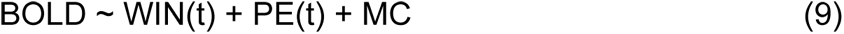

In Equation 9, WIN(t) indicates whether the subject obtained positive feedback in the trial, and PE represents the prediction error of the received feedback calculated based on the behavioral model.

Structural Image processing was conducted using the first two steps of the Human Connectome Project (HCP) pipeline, detailed elsewhere.^31^ Briefly, the “PreFreeSurfer” phase generated an undistorted native structural volume space, aligned T1-weighted (T1w) and T2-weighted (T2w) images, corrected bias fields derived from both images, and registered the native space to the common MNI coordinate space using a rigid affine registration, and them, a non-linear registration to standard space. Utilizing FreeSurfer version 6, the “FreeSurfer” stage conducted volume segmentation and cortical surface reconstructions, including delineation of the “white” (gray/white matter boundary) and “pial” (gray/cerebrospinal fluid [CSF] boundary) surfaces, followed by registration to a common template (fsaverage). Whole-brain analyses were conducted across cortical surface vertices, and corrections were applied to minimize the Type II errors.^32^ These analyses utilized Surfstat, a MATLAB toolbox (available at http://www.math.mcgill.ca/keith/surfstat/). Corrections based on random field theory (RFT) were applied (cluster-corrected p < 0.05, cluster threshold detection, CTD, z > 3.1) to address multiple comparisons (see an example in^33^).

Diffusion imaging processing was computed using DSI Studio software (http://dsi-studio.labsolver.org/), which performs Eddy current and motion correction and calculates the DTI model. It also extracts diffusion measures such as fractional anisotropy (FA), mean diffusivity, axial diffusivity, and radial diffusivity. Deterministic tractography algorithm^34^ was applied using the following parameters: angular threshold=60°, step size= 1mm, minimum length= 30mm, maximum length= 250mm, smoothing= 0.5, and QA threshold=0. The fiber segmentation was performed using a DWM bundle atlas,^35^ which consists of 36 known bundles, and a SWM bundle atlas,^36^ composed of 525 short bundles, from which the 209 most stable bundles for deterministic tractography were selected. An automatic segmentation algorithm was used based on the maximum Euclidean distance between the corresponding points of two fibers.^37^

Voxelwise statistical analysis of the diffusion data was conducted using TBSS.^38^ Initially, FA data from all subjects were aligned to the 1 × 1 × 1 mm³ FMRIB58_FA standard space atlas via nonlinear registration. Subsequently, a mean FA image was generated from the aligned FA images and then thinned to form a skeletonized mean FA, representing the central axes of all tracts common to all subjects in the analysis. The mean FA skeleton was thresholder to FA ≥ 0.2 to include major white matter pathways, excluding peripheral tracts and cortical gray matter. Each subject’s aligned FA data were projected onto the skeleton by searching perpendicularly from the skeleton for maximum FA values in each individual’s FA maps. Statistical comparisons of FA maps were subsequently confined to voxels within the white matter skeleton. We utilized Randomise (FSL program package) to conduct voxelwise statistics on the skeletonized FA data. A multiple linear regression model used FA data as the dependent variable. The independent variables are shown in Figure 2A. Permutation-based testing (5000 permutations) and statistical inference were performed with correction for multiple comparisons conducted through threshold-free cluster enhancement (TFCE).

**Figure 2.**
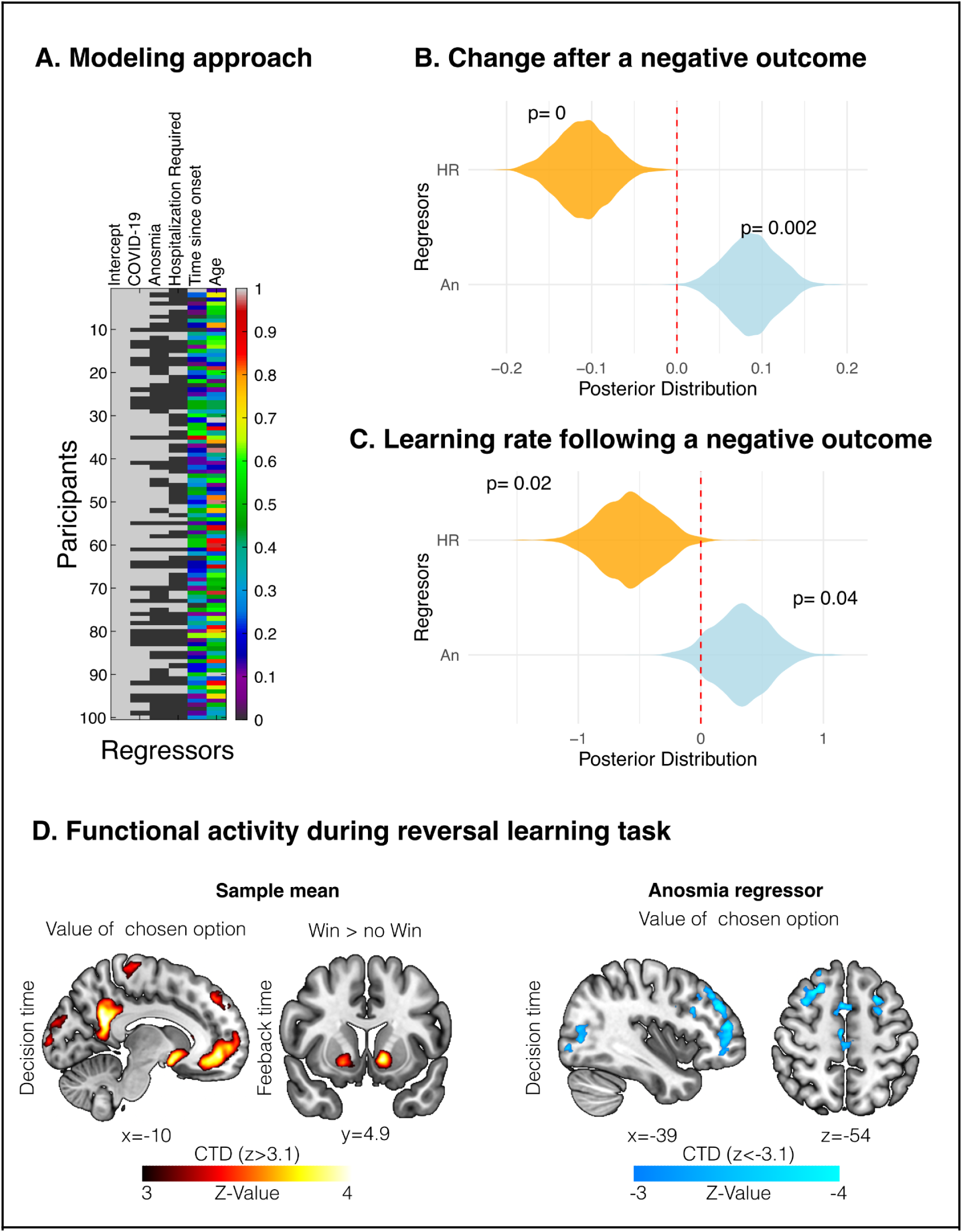
Behavioral and functional results. **A.** Model applied to behavioral and brain data. **B-C.** Behavioral data from Reversal Learning Task. **B.** Regressor effects over the rate of option change after a negative outcome during shift periods. **C.** Regressor effects over learning rate following a negative outcome. **D.** BOLD activity during the Reversal Learning Task. The left panel shows the global effect of the task. The right panel indicates the negative effect of the Anosmia regressor. HR: Hospitalization required; An: Anosmia; CTD: cluster-threshold detection.

### Statistical Analyses

We employed general linear models (GLM) using the R software to analyze the sample’s demographic characteristics. The same modeling approach, as depicted in Figure 2A, was utilized to compare all parameters between patients and controls and between clinical factors (anosmia and hospitalization requirement). In this approach, target measurements were used as dependent variables, while COVID-19 diagnosis, anosmia during the acute episode, and hospitalization requirement during the acute episode were utilized as independent variables. Given the statistical differences observed in some demographic variables between COVID-19 patients who required hospitalization and those who did not (see below), we augmented the model by including age and the time between diagnosis and evaluation as control regressors. Residual analysis was applied to assess the validity of the fitted model and verify underlying assumptions using the observed data. We used normalized parameters to indicate effect sizes per regressors in regression analyses, displaying their respective 95% confidence intervals. All key findings of behavioral results derived from GLM were replicated using Bayesian estimation. The results were presented in the figures depicting the posterior distribution of the parameters and the p_MCMC_ derived from this analysis (p_MCMC_ is a p-value derived by comparing the posterior distributions of the estimated parameters sampled via Markov Chain Monte Carlo; see section “Cognitive modeling” for details). Additionally, for testing behavioral indicators in the Reversal Learning Task, we employed non-parametric statistics (since these indicators generally have a non-normal distribution^39–43^) and their respective effect size measures, together with mixed linear models.

## Results

### Demography

We aimed to evaluate for cognitive, structural, and functional alteration in patients recovering from COVID-19 and how this alteration depends on the clinical profile of the patients. Two clinical factors were assessed: Anosmia (**An**) during the acute episode as a potential marker for affection other than respiratory inflammation, and hospitalization (**HR**) during the acute episode to indicate the severity of respiratory symptoms. Using linear modeling, we observed variations in age and the time elapsed between diagnosis and the first evaluation among patients exhibiting these factors. Concerning age, there were no significant differences observed between patients with and without COVID-19 (age in z-score, beta=0.05, se=0.2, t=0.2, p=0.8) or between patients with and without anosmia (beta=-0.05, se=0.2, t=-0.2, p=0.8). However, patients with COVID-19 who required hospitalization were older than those with COVID-19 who did not require hospitalization (beta=0.7, se=0.2, t=3.4, r=0.35 [0.15 0.55], p=0.0008). Additionally, patients with COVID-19 requiring hospitalization presented a longer time interval between diagnosis and evaluation (time in z-score, beta=0.6, se=0.2, t=3.1, r=0.32 [0.12 0.52], p=0.002). Consequently, age and time between diagnosis and evaluation were used as control regressors in all analyses comparing clinical factors, as indicated in Figure 2A.

### Persistent symptoms

All patients were queried about persistent post-COVID symptoms during evaluation.^44^ Twenty-two patients diagnosed with COVID-19 reported experiencing some degree of attention and memory issues, which persisted at the time of evaluation. The frequency of these reported cognitive symptoms did not show modulation by clinical factors (linear model dof=93, Anosmia: beta=0.05, se= 0.09, t=0.5, r=0.06 [-0.17 0.29], p=0.6; Hospitalization required: beta=0.08, se=0.09, r=0.09 [-0.13 0.32], t=0.8, p=0.4). Additionally, seven patients reported cephalea, and six reported fatigue. Only four patients reported persistent olfactory alteration post-acute episodes. Patients underwent screening for olfactory deficits due to SARS-CoV-2 using the KOR test;^26^ of these, six (out of 43 patients with anosmia during the acute episode) met the criteria for persistent olfactory deficit. Despite this, when evaluating the KOR test scores with the model (Figure 2A), no group differences were observed (linear model dof=93, Anosmia: beta=-0.39, se=0.24, t=-1.6, r=-0.19 [-0.44 0.05], p=0.11; Hospitalization required: beta=-0.14, se=0.24, t=-0.5, r=-0.07 [-0.30 0.17], p=0.5). Functional capacity was also evaluated using 6MWT.^27^ We did not find differences in this score between groups (linear model dof=93, An beta=-0.003, se=0.02, t=-0.16, r=0.06 [-0.17 0.29], p= 0.8; HR beta=0.007, se=0.02, t=0.3, r=0.09 [-0.13 0.32], p=0.7).

### Psychological assessment

Patients were evaluated using cognitive screening. ACE-III evaluation showed that the sample had a mean score of 92 with no differences between groups (linear model, df=93, COVID-19 diagnosis beta=-1.9, se=3.0, t=-0.6, r=-0.08 [-0.33 0.17], p=0.5; Anosmia beta=3.2, se=2.6, t=1.1, r=0.15 [-0.10 0.39], p=0.2; Hospitalization required beta= –3.0, se=2.7, t=-1.0,r=-0.13 [-0.3 0.11], p=0.2). In the same way, IFS-Ch frontal screening evaluation showed a mean of 21.8 with no differences between groups (COVID-19 diagnosis beta=1.2, se=1.1, t=1.1, r= 0.13 [-0.11 0.38], p=0.2; Anosmia beta=0.5, se=1.0, t=0.4, r=0.06 [-0.18 0.29], p=0.6; Hospitalization required beta= –3.2, se=1.0, t=-0.3, r=-0.04 [-0.26 0.19], p=0.7). We found similar results when analyzing the PHQ-9 (COVID-19 diagnosis beta=2.0, se=1.4, t=1.4, r=0.18 [-0.07 0.43], p=0.15; Anosmia beta=-0.9, se=1.2, t=-0.7, r=-0.09 [-0.34 0.15], p=0.4; Hospitalization required beta= –0.5, se=1.2, t=-0.4, r=-0.05 [-0.29 0.19], p=0.6), and GAD-7 screenings (COVID-19 diagnosis beta=2.5, se=1.4, t=1.8, r=0.22 [-0.02 0.47], p=0.07; Anosmia beta=0.2, se=1.2, t=0.1, r=0.02 [-0.22 0.26], p=0.8; Hospitalization required beta= –0.07, se=1.2, t=-0.06, r=-.007 [-0.24 0.23], p=0.9).

### Behavioral Task

Initially, we assessed whether participants adapted their behavior during the game (Figure 1). For this purpose, we analyzed three phases during the game: a phase we labeled as ‘Shift,’ which encompasses the five trials following the programmed probability change; a ‘Pre-Shift’ phase, comprising the last five trials of the initial stable phase of each game, and a final ‘Post-Shift’ phase, representing the final five trials of each game (corresponding to the conclusion of the second stable phase, see Figure 1C).

All participants decreased their earnings during the Shift phase, but increased them in the Post-Shift phase, reflecting learning and adaptation (Friedman test, stat=44.8, df=2, Kendall W = 0.22, p=2e-10; mixed model over single trials, b=-0.14, se=-0.01, t=-10.29, r=-0.11 [-0.13 –0.09], p=2e-16, Figure 1B). Subsequently, we investigated an indicator of the strategies individuals employ during transitions. To do so, we assessed the rate of alternative change following a negative outcome. An exceedingly low value in this indicator suggests a tendency for individuals to uphold the value of the chosen option after experiencing negative outcomes, a phenomenon known as perseverative decision-making^45^. Conversely, exceedingly high values in this indicator may signify impulsive shifts or the tendency to alter one’s choice immediately following an error without updating the value. This indicator decreases in value during the Shift compared to the Pre-Shift phase, reflecting the tendency to accumulate more evidence before shifting from the previously advantageous option (linear model, averaged data: beta=-0.04, se=0.01, t=-2.6, r=-0.11 [-0.19 –0.03], p=0.008; mixed-effects logistic model over single trials: beta=-0.24, se=0.05, t=-4.4, r=-0.11 [-0.15 –0.06], p=9e-6). These initial analyses indicate that the entire sample exhibited the expected behavior in the task, adapting their decisions after a shift with a cost in the transition.

Next, we applied the strategy indicator during the Shift phase to investigate potential differences among groups using the model described in Figure 2A. We observed that the clinical characteristics of COVID-19 patients differentially influenced the strategy indicator. The diagnosis of COVID-19 did not significantly impact the indicator (linear model: beta=0.01, se=0.03, t=0.5, r=0.04 [-0.12 0.21], p=0.6); however, patients requiring hospitalization exhibited a decrease in this parameter (linear model: beta=-0.1, se=0.03, t=-3.4, r=-0.28 [-0.44 –0.12], p=0.0007, similar results from Bayesian estimation shown in Figure 2B). In contrast, patients presenting anosmia demonstrated an increase in this parameter (linear model: beta=0.08, se=0.03, t=2.8, r=0.24 [0.07 0.40], p=0.004, similar results from Bayesian estimation shown in Figure 2B). None of this modulation occurred in the other phase of the task (ps>0.1). This strategic modulation significantly impacted total earnings, leading to higher earnings among patients with anosmia (linear model, beta=0.02, se=0.01, t=2.43, r=0.10 [0.02 0.19], p=0.015).

Then, we test if this behavioral modulation is related to specific cognitive computation. We fitted a cognitive model of participants’ responses using prospect theory and a Rescorla-Wagner algorithm to estimate the individual learning of the probability of each desk. We used a different learning rate estimated following a win and a no-win. Based on the preceding results, we tested if the clinical condition of hospitalization and anosmia modulated the differences between the learning rates. We found a similar pattern to the prior results: COVID-19 diagnosis per se did not affect the learning rate (one tail test, linear model df=90, b=0.1, se=0.2, t=0.4, r=0.1 [-0.14 0.34], p=0.3), Hospitalization generated a decrease in the learning rate after negative outcome (b=-0.51, se=0.24, t=-2.1, r=-0.27 [-0.50 –0.04], p=0.018, similar results from Bayesian estimation shown in Figure 2C), and Anosmia presents a tendency to increases learning rate (b=0.4, se=0.2, t=1.8, r=0.2 [0 0.43], p=0.04, similar results from Bayesian estimation shown in Figure 2C).

In summary, participants adjusted to the changing probabilities, resulting in increased earnings following the decreases caused by the shift in probability. A behavioral indicator shows participants’ ability to employ different strategies during reversals. Clinical characteristics of COVID-19-recovered patients influenced this indicator, with hospitalized patients decreasing and anosmic patients increasing, impacting total earnings. When testing specific cognitive computations, the modulation due to hospitalization affected the individual learning rate.

### Brain Functional Differences

We evaluated the BOLD signal of the participants while they engaged in the Reversal Learning Task. Cognitive modeling was used to estimate the utility of the chosen option (see Materials and Methods). During the feedback period, we contrasted wins and no wins.

Initially, we assessed the consistent activity across the entire sample to identify the activity associated with value and feedback as classically described in this type of task. We found that during the decision-making process, the value of the chosen option correlated with an extensive frontal-parietal-striatal network, consistent with the literature, including ventromedial prefrontal, medial parietal, and striatal regions.^29,46^ Conversely, during feedback, we observed that the contrast between win and non-win revealed activity in the ventral striatum, consistent with prior research.^29^ Subsequently, we assessed the modulation of clinical parameters on BOLD activity. COVID-19 diagnosis and hospitalization required regressors did not show modulation in decision-related or feedback-related activity. However, the regressor associated with anosmia negatively modulated the BOLD signal during decision-making in a network that includes lateral prefrontal, medial frontal, and left temporoparietal regions.

### Brain Structural Differences

#### Cortical thickness

The gray and white matter were segmented using T1w and T2w images. Cortical thickness was analyzed using the specified model in the methods (Figure 2A). We found that neither the COVID-19 diagnostic regressor nor the hospitalization requirement showed significant modulation in cortical thickness. However, the anosmia correlated with a thinning of the cortical thickness in parietal areas (Figure 3A).

**Figure 3.**
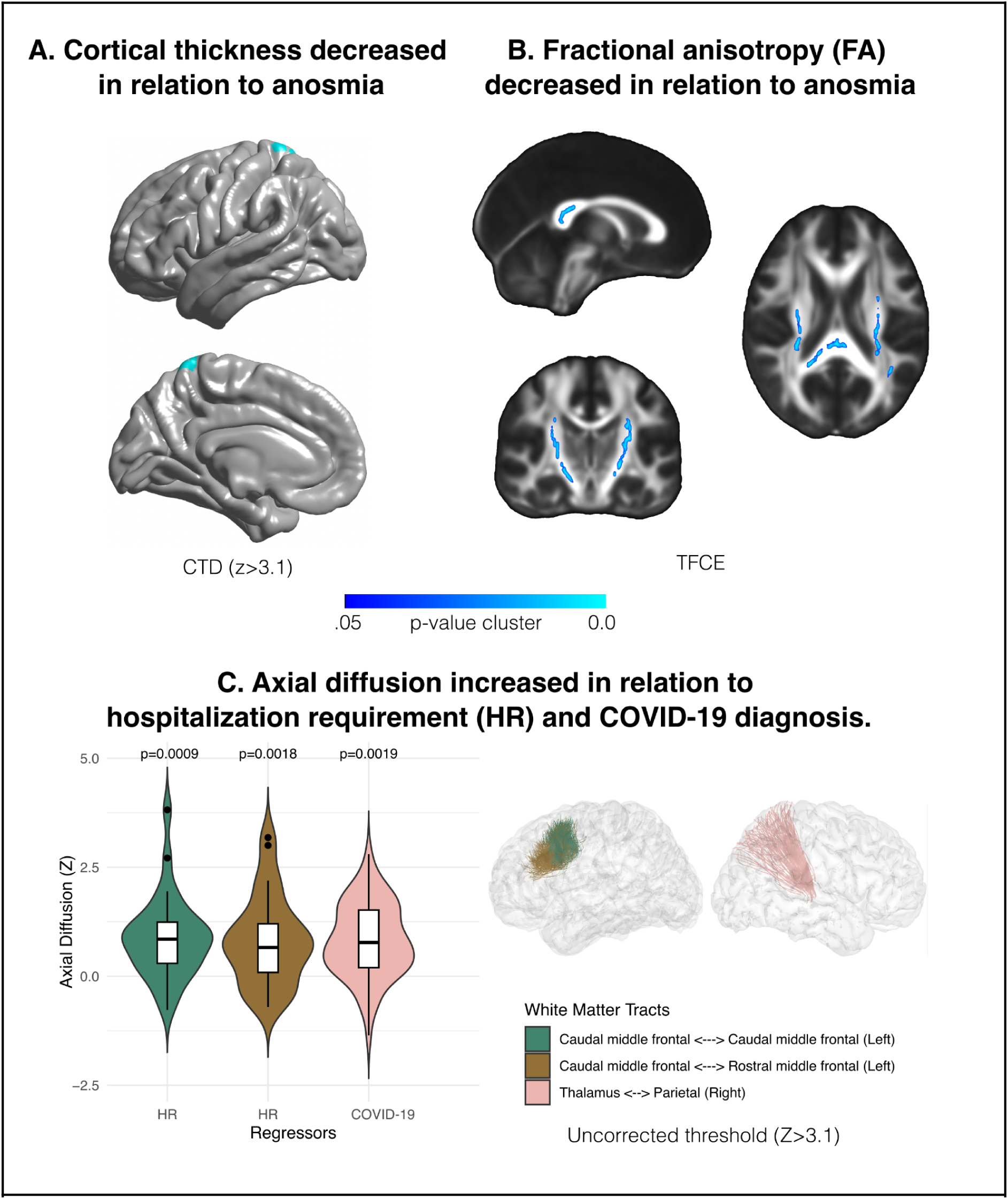
Brain structural results. **A.** The anosmia regressor effect over the cortical thickness. **B.** The anosmia regressor effect over the fraction of anisotropy in a whole-brain analysis of white matter integrity. **C.** Regressor effects over axial diffusivity measuring in segmented white matter tracts. HR: hospitalization required, CTD: cluster-threshold detection, TFCE: threshold-free cluster enhancement.

#### White matter integrity

The integrity of the white matter was assessed through diffusion images. First, we conducted a whole-brain analysis, evaluating changes in the fractional anisotropy (FA). Statistical modulations were calculated using the specified model outlined in the methods (Figure 2A), and cluster-based statistics were performed using TFCE. Anosmia was the only regressor with significant modulation, demonstrating decreased FA (Figure 3B). The main tracts involved in the affected areas were the corticospinal tract, arcuate fasciculus, inferior frontal-occipital fasciculus, thalamus-parietal fasciculus, thalamus-occipital fasciculus, and posterior corpus callosum.

Next, we conducted statistical analyses for individual tracts. Long and short fibers were segmented using deterministic tractography. Various diffusion measures were evaluated to assess the integrity of each tract (FA, radial diffusion, axial diffusion, and mean diffusion). Each tract was evaluated using the model specified in the methods. The analysis revealed that no modulation survived multiple comparisons (Bonferroni correction). However, when applying an uncorrected threshold (Z > 3.1, commonly used for cluster detection in whole-brain functional and structural imaging studies), it was observed that frontal and parietal fascicles exhibited an increase in axial and mean diffusion, indicating a disruption in white matter integrity. This white matter integrity disruption correlated with the hospitalization requirement and COVID-19 diagnosis (Figure 2C).

## Discussion

The results of our study give new insight into the cognitive, structural, and functional alterations observed in patients recovering from COVID-19, with a particular focus on how these alterations relate to the patient’s clinical profiles.We assessed two key clinical factors, anosmia during the acute episode and the requirement for hospitalization, as proxies for potential markers of neurological involvement and disease severity, respectively.

Previous reports have found a high prevalence of smell or taste dysfunction in long-term COVID-19, with a percentage of patients being functionally anosmic even one year after SARS-CoV-2 infection.^47^ Those patients with persistent COVID-19-related anosmia showed altered olfactory network connectivity correlated with hyposmia severity and neuropsychological performance.^48^ Also, memory and mood disturbances in long-COVID patients with mild or moderate disease correlated with hyposmia or ageusia.^49^ Our findings build upon prior research by indicating that the presence of anosmia during the acute phase, despite transient symptoms, indicates possible brain alterations. Only six patients present indicators of persistent olfactory deficit; thus, our results are not due to actual deficit. Hence, anosmia can serve as both a potential marker of virus-induced damage to neuronal tissues and a marker for individuals susceptible to brain damage.

Regarding cognitive assessment, our findings revealed no significant differences in cognitive performance between groups based on COVID-19 diagnosis, anosmia, or hospitalization requirements. Most of the research that has identified manifest cognitive deficits typically involves more severe respiratory cases of COVID-19^50–52^ or those who present neurological symptoms during the acute episode.^9^ Additionally, these patterns of results are more likely to be found when focusing on a subset of the population that likely represents a more susceptible group, such as those individuals who report subjective cognitive symptoms persisting for more than a month after infection.^9^ A study using web-based assessments over a large sample of subjects found that individuals recovering from COVID-19 exhibited alterations in cognitive tests, with more pronounced effects observed in those who were hospitalized.^53^ However, significant alterations were also evident in those who did not require hospitalization.^53^ While no differences were observed in our cognitive screening evaluation, intriguing patterns emerged in behavioral task performance. All participants displayed adaptive behavior during the game, as evidenced by increased earnings after the shift phase. Particularly, the distinct decision-making strategy observed in patients with anosmia was characterized by more impulsive option shifts during reversals. In our task’s context, marked by high uncertainty and volatility, this resulted in higher total earnings than other groups. Conversely, patients requiring hospitalization exhibited an opposing pattern, showing a decrease in the strategy indicator and more perseverative decision-making. A prior study has reported decision-making alterations in patients who recovered from COVID-19.^54^ Our results provide further insight into this previous evidence, highlighting the existence of distinct patterns of behavioral alteration, which are related to the clinical profile and may also reflect different physiological mechanisms.

Corresponding patterns in brain activity accompanied these behavioral differences. Anosmia was associated with decreased BOLD signal during decision-making in specific brain regions, including lateral prefrontal, medial frontal, and left temporoparietal regions. The change corresponds to a decrease in value-related activity during decision-making. Thus, a weak value signal can generate a more impulsive option change after negative results. Most studies investigating functional brain activity in patients recovered from COVID-19 have been conducted using resting-state methods. Alterations in signal properties and functional connectivity have been found in COVID-19 patients,^55,56^ and have been associated with hospitalization,^57^ and persistent symptoms such as headaches,^58^ persistent olfactory dysfunction ^48,59,60^ and other behavioral markers such as working memory performance,^61,62^ anxiety,^63^ psychotic-like experiences in adolescents,^64^ and post-traumatic stress symptoms.^65^ Also, some studies found no functional alteration in recovered COVID-19 patients.^66^ Taking this evidence into account, our results reveal a more nuanced landscape concerning the functional alterations following COVID-19 infections. While a clear pattern emerges regarding persistent symptoms reflected in functional brain activity, an additional pattern is likely an indicator of subtle brain damage or susceptibility among patients who exhibit alterations without clearly manifesting cognitive or persistent symptoms. A history of anosmia during infection may serve as a risk factor in this latter group.

Our structural results support this interpretation, where the main findings are related to a history of anosmia and not hospitalization. Specifically, cortical thickness analysis revealed thinning in parietal areas among patients with anosmia, indicating structural alterations in these regions. Moreover, analysis of white matter integrity demonstrated significant alterations in patients with anosmia, as evidenced by decreased fractional anisotropy in various white matter tracts, including the corticospinal tract and arcuate fasciculus. Studies of large samples of patient recovery from COVID-19 have shown alteration of these brain structural parameters.^1,2^ These studies indicate that subtle brain changes are present even in non-severe cases of COVID-19 without manifest cognitive alterations.^1,2^ However, research has failed to find a specific marker of neuroinflammation,^67^ or structural alterations were not found in other samples.^48,62^ As with functional alterations, the severity of the acute episode and the presence of persistent symptoms are more closely related to structural alterations, primarily in white matter integrity. ^66,68^

Overall, our results highlight the importance of a history of anosmia in relation to the presence of brain alterations. Given the substantial number of patients recovering from COVID-19 worldwide, it is crucial to identify risk factors associated with potential brain damage. In this context, a history of anosmia can be a useful criterion for prioritizing deeper follow-up of these patients. To what extent does anosmia reflect a specific neuropathology of brain damage derived from COVID-19, or is it a marker of patient susceptibility to different neuropathological mechanisms? This remains an open question that necessitates further research. Some evidence suggests that anosmia may reflect more on a patient’s susceptibility to brain damage rather than being part of the neuropathology of brain damage. Most of our patients experience anosmia as a transient symptom during acute episodes yet still exhibit brain alteration signatures. Moreover, alteration in the central olfactory system related to persistent olfactory deficits are less severe in patients who recovered from COVID-19 than those with other causes of persistent olfactory deficits.^69^ Interestingly, studies on the presence of anosmia due to different SARS-CoV-2 variants and genetic associations have shown that these symptoms are associated with the UGT2A1/UGT2A2 locus, which encodes an enzyme expressed in the olfactory epithelium and the brain.^20^ How this finding reflects the mechanism of the association between anosmia and brain alteration requires further investigation.

Our study provides valuable insights into the complex interplay between clinical factors, cognitive performance, and brain alterations in COVID-19 recovery patients. These findings contribute to our understanding of the neurological consequences of COVID-19 and underscore the importance of early detection and intervention in at-risk populations. Additional research is necessary to clarify the underlying mechanisms and identify potential therapeutic targets for mitigating long-term neurological sequelae in COVID-19 patients.

## Data and code availability

All data will be available in the OpenNeuro repository before publication. All codes will be available in the GitHub repository (https://github.com/neurocics/Kausell_FigueroaVargas_COVID) before publication.

## Acknowledgments

We thank Karen Czischke for her clinical assistance.

## Funding

This work was supported by Universidad del Desarrollo, Proyecto interno 2020 23400175 to LK, Clínica Alemana de Santiago Proyecto de investigación ID 1033 to XS, Agencia Nacional de Investigación y Desarrollo de Chile (ANID), FONDECYT (1211227 to PB, LK, AF-V and PS-I; 1190513 to FZ and PB; 1221837 to PMV; 11230607 to PS-I), FONDEQUIP EQM150076, ANID-Basal Project FB0008 (AC3E) to PG.

## Competing interests

The authors report no competing interests.

## Declaration of generative AI and AI-assisted technologies in the writing process

During the preparation of this work the authors used ChatGTP 3.5 in order to improve the readability and language of the manuscript. After using this tool, the authors reviewed and edited the content as needed and take full responsibility for the content of the published article.

## Notes

### Competing Interest Statement

The authors have declared no competing interest.

### Funding Statement

This work was supported by Universidad del Desarrollo, Proyecto interno 2020 23400175 to LK, Clinica Alemana de Santiago Proyecto de investigacion ID 1033 to XS, Agencia Nacional de Investigacion y Desarrollo de Chile (ANID), FONDECYT (1211227 to PB, LK, AF-V and PS-I; 1190513 to FZ and PB; 1221837 to PMV; 11230607 to PS-I), FONDEQUIP EQM150076, ANID-Basal Project FB0008 (AC3E) to PG.

### Author Declarations

All participants gave their informed consent, and all experimental procedures were approved by the Ethics Committee of Clinica Alemana – Universidad del Desarrollo, Chile (Folio 2020-102).

